# Dengue in western Uganda: A prospective cohort of children presenting with undifferentiated febrile illness

**DOI:** 10.1101/2020.08.21.20179002

**Authors:** Ross M. Boyce, Matthew Collins, Rabbison Muhindo, Regina Nakakande, Emily J. Ciccone, Samantha Grounds, Daniel Espinoza, Yerun Zhu, Michael Matte, Moses Ntaro, Dan Nyehangane, Jonathan J. Juliano, Edgar M. Mulogo

**Affiliations:** Division of Infectious Diseases, University of North Carolina at Chapel Hill, Chapel Hill, North Carolina, 27599, USA; Department of Community Health, Faculty of Medicine, Mbarara University of Science & Technology, Mbarara, Uganda; Division of Infectious Diseases, Emory University, Atlanta, Georgia, 30322, USA; College of Arts and Sciences, University of North Carolina at Chapel Hill, Chapel Hill, North Carolina, 27599, USA; Epicentre Mbarara Research Centre, Mbarara, Uganda

**Keywords:** Dengue, fever, arbovirus, epidemiology, Uganda

## Abstract

**Background:** The spatial distribution and burden of dengue in sub-Saharan Africa remains highly uncertain, despite high levels of ecological suitability. The goal of this study was to describe the epidemiology of dengue among a cohort of febrile children presenting to outpatient facilities located in areas of western Uganda with differing levels of urbanicity and malaria transmission intensity.

**Methods:** Eligible children were first screened for malaria using rapid diagnostic tests. Children with a negative malaria result were tested for dengue using a combination NS1/IgM/IgG rapid test (SD Bioline Dengue Duo). Confirmatory testing by RT-PCR was performed in a subset of participants. Antigen-capture ELISA was performed to estimate seroprevalence.

**Results:** Only 6 of 1,416 (0.42%) children had a positive dengue rapid test, while none of the RT-PCR results were positive. ELISA testing demonstrated reactive IgG antibodies in 28 (2.2%) participants with the highest prevalence seen at the urban site in Mbarara (19 of 392, 4.9%, p< 0.001).

**Conclusions:** Overall, these findings suggest that dengue, while present, is an uncommon cause of non-malarial, pediatric febrile illness in western Uganda. Further investigation into the eocological factors that sustain low-level transmission in urban settings are urgently needed to reduce the risk of epidemics.

## Background

Dengue is a mosquito-borne viral disease that is estimated to cause upwards of 400 million infections each year [1, 2]. While more than half of the world’s population is thought to be at risk, the global distribution and burden of dengue remains highly uncertain [3]. Nowhere is this epidemiological uncertainty more pronounced than in Africa, where the requisite laboratory infrastructure to distinguish dengue from other causes of febrile illness is not routinely available [4]. Despite limited data, there is reasonable consensus for the existence of endemic dengue transmission in many countries, with modeling frameworks suggesting that Africa’s disease burden may be similar to that of other high transmission areas, such as the Americas [1].

The indirect evidence supporting endemic dengue transmission in Uganda is relatively strong. There are favorable precipitation and temperature conditions, the *Aedes aegypti* mosquito – the primary vector of dengue, along with yellow fever and Rift Valley fever – is ubiquitous in Uganda. Dengue outbreaks and transmission have been documented in neighboring countries, and cases have been reported among travelers returning from Uganda [1, 5, 6]. Other factors favoring the likelihood of dengue transmission in Uganda include an increasingly globalized economy and rapid urbanization [7, 8].

While the majority of dengue infections result in either asymptomatic or mild, self-limited clinical disease, the potential health system and economic impact of dengue transmission in a malaria-endemic country like Uganda is significant but also not well understood. Given the non-specific symptoms and lack of available diagnostic tools, dengue infections may frequently be misdiagnosed and empirically treated as malaria [6]. This likely contributes to (i) over-estimation of malaria transmission, (ii) over-use of artemisinin combination therapies (ACT), perhaps accelerating to the development of resistance, and (iii) inadequate resourcing of dengue surveillance and control measures, which generally do not overlap with malaria control strategies such as insecticide-treated nets (ITN) [9].

Defining the burden of dengue in countries like Uganda is critically important, especially as new tools such as vaccines become more widely available [10, 11]. Therefore, the overarching aim of this study was to describe the epidemiology of dengue among a cohort of children presenting to outpatient public health facilities in western Uganda with acute febrile illness.

## Methods

### Study Setting

The study was conducted at two sites in the Kasese District and one site in the Mbarara District of Southwest Uganda (**Figure 1**). These facilities were purposefully selected to examine dengue as a potential cause of non-malarial febrile illness across different ecological settings and in areas of varying malaria transmission intensity (**Table 1**). There are no government-provided diagnostic tools or standardized treatment protocols for dengue fever at public health facilities in Uganda [12].

**Figure 1:**
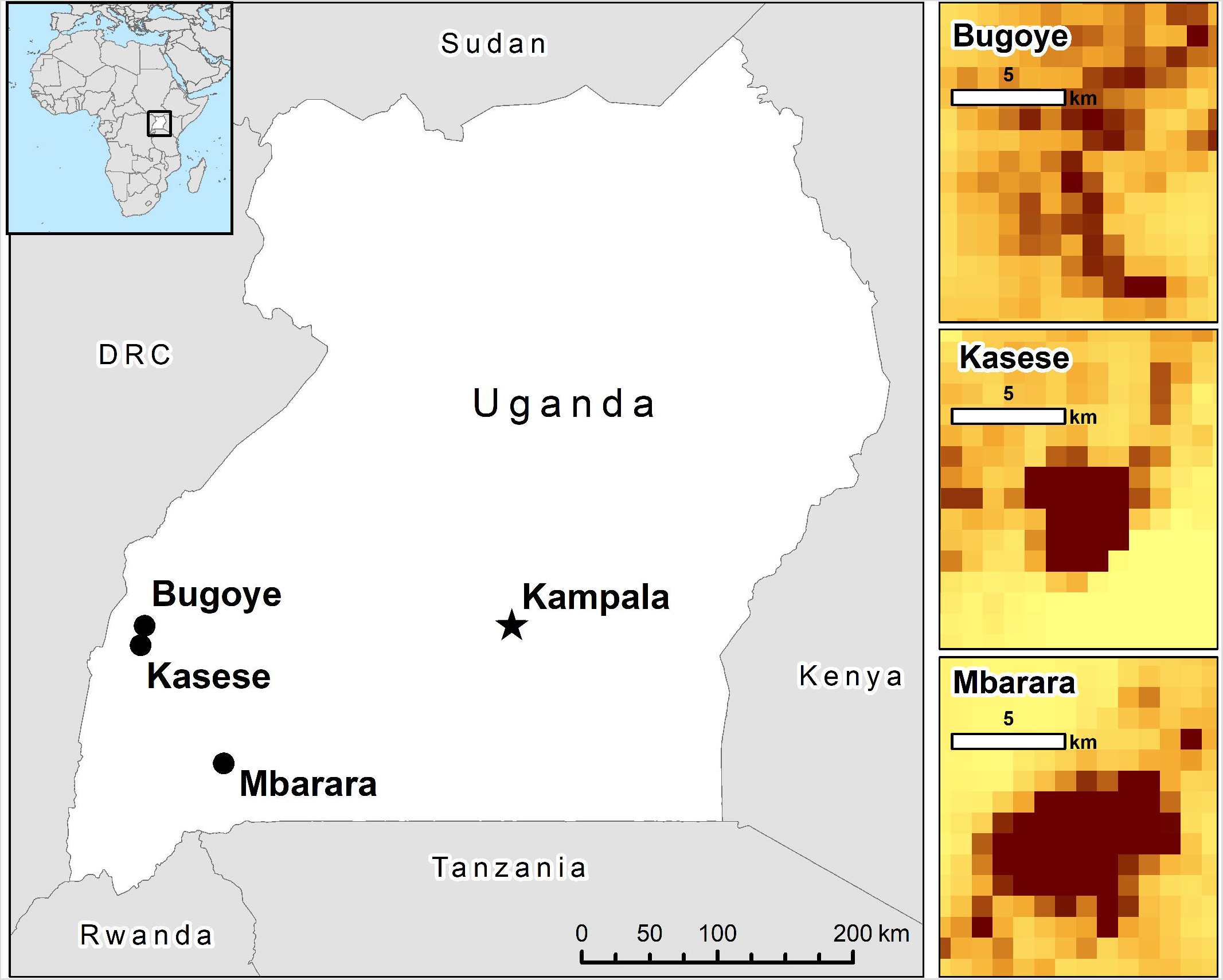
Map showing location of study sites and population density of each location.

**Table 1:** Description of clinical sites

| Study Site | Location | District | GPS | Elevation | Setting | Facility Type | Referral Base | Monthly Clinic Visits | Malaria Prevalence* |
| --- | --- | --- | --- | --- | --- | --- | --- | --- | --- |
| Bugoye Health Centre III | Bugoye Village | Kasese | 0° 30' N<br>30° 1' E | 1,242m | Rural | Public Health Centre III | 50,000 | 800 | 17.6% |
| Kasese Health Centre III | Kasese Town | Kasese | 0° 17' N<br>30° 7' E | 980m | Semi-Urban | Public Health Center III | 400,000 | 1,200 | 17.6% |
| Mbarara Regional Referral Hospital | Mbarara Town | Mbarara | 0° 36' S<br>30° 4' E | 1,442m | Urban | Public Teaching Hospital | 2.5 million | 1,600 | 5.7% |

\*Regional estimates from 2014-15 Uganda Malaria Indicator Survey

### Study Design

The study was a prospective, observational cohort design, enrolling malaria-negative, febrile children from November 2017 to June 2018. Children (age < 18 years) presenting to one of three outpatient clinics with documented fever (axillary temperature ≥38° celsius [C]) or a reported history of fever within the last 7 days were eligible for inclusion. Children presenting without a parent or guardian to provide consent were excluded. After consent was provided, study personnel recorded demographic information, vital signs, and administered a brief health questionnaire.

Laboratory staff performed a malaria rapid diagnostic test (mRDT) (SD Bioline Malaria Ag P.f, Standard Diagnostics, Republic of Korea) and collected dried blood spots (DBS) on filter paper (Whatman, Chicago, IL). DBS sampling was chosen as a pragmatic but effective option for sampling children across the different field conditions at our study sites [13]. The reliability of measuring antibodies eluted from DBS has been validated several times, [14, 15] and we and others have used DBS-based serology to study the epidemiology of arboviruses in both standard ELISA as well as multiplex platforms [16-18]. Children with a positive mRDT result were excluded from further participation.

Children with a negative mRDT result subsequently underwent testing with a dengue rapid diagnostic test (dRDT) (SD Bioline Dengue Duo, Standard Diagnostics Korea). Participants at the Mbarara Regional Referral Hospital outpatient clinic, where a reliable cold chain was available, had 5 mL of venous rather than capillary blood drawn into serum serparator tubes for further testing in addition to the dRDT.

Results of the dRDT were provided to the responsible clinician for the purposes of counseling and case management. Treatment and disposition (i.e. admission versus discharge) plans, as determined by the clinician, were recorded. DBS were stored at room temperature under desiccation and serum samples were stored at –20 °C until analysis.

### Laboratory Methods

All RDTs for the study were obtained directly from the manufacturer, stored in the original packaging at room temperature, and used in accordance with the manufacturer’s instructions prior to the expiration date. Further description of specific procedures is provided below.

### Elution of dried blood spots for serologic analysis

Plasma proteins were eluted from DBS as previously described [16–18]. One 6-mm hole punch of each DBS was placed in a 1.5 mL Eppendorf tube with 300 μL of phosphate-buffered saline, and rotated for 2 hour at 37°C. This yielded an eluate that is equivalent to a 1:40 plasma dilution. Eppendorf tubes were centrifuged then eluate transferred to a new tube. Eluate was heat inactivated for 30 minutes in a 56°C water bath. The samples were centrifuged again to pellet proteinaceous debris and the supernatant was transferred to a new tube and stored at 4°C for up to 1 week or at –20°C until use.

### Antigen capture IgG ELISA

Binding IgG to DENV or ZIKV was measured by antigen capture ELISA as previously described [19]. Briefly, DENV antigen (an equal volume mixture of supernatant from each of the four DENV serotypes cultured in C6/36 cells) was captured by the anti-E protein mouse mAb 4G2 [20]. Plates were blocked with 3% nonfat dry milk, and incubated with DBS eluate at 37oC for 1 hour, and binding was detected with an alkaline phosphatase-conjugated goat anti-human IgG secondary Ab and *p*-nitrophenyl phosphate substrate. Absorbance at 405 nm (optical density, OD) was measured by spectrophotometry on a plate reader. ELISA data are reported as OD values that are the average of technical replicates. The average OD for technical replicates using DBS eluate obtained from flavivirus-naïve individuals (NHS) served as the negative control in ELISA assays. The cut off for positivity was calculated for each plate as the average OD of NHS = standard deviations + 0.1 [16–18].

### IgM ELISA

Testing for anti-DENV IgM was only performed on participant samples with either a positive dRDT or IgG ELISA per CDC MAC ELISA protocol after titrating individual reagents per instructions [21]. DBS eluate was tested at 1:40 dilution. DENV1–4 antigen was the same as for IgG ELISA above. Plates were washed 3 times between each step. The Enhanced K-Blue TMB substrate reaction was stopped after 30 minutes by addition of 50 μL 1N HCl. Optical density (OD) of each well was determined within 5 minutes at 450nm.

### Neutralization Assays

Neutralization titers were determined by 96-well microFRNT [22, 23]. Due to limited sample availability from DBS, an abbreviated neutralization assay format was used (eFRNT). DBS eluates were run in singleton over four 4-fold dilutions. Serial dilutions of DBS eluate were mixed with approximately 75–100 focus-forming units of virus in DMEM with 2% FBS. The virus-antibody mixtures were incubated for 1 hour at 37°C and then transferred to a monolayer of Vero cells for infection for 2 hours at 37°C. OptiMEM overlay media supplemented with 2% FBS and 5g (1%) Carboxymethylcellulose was then added, and cultures were incubated for 48 hours (DENV2 and DENV4) or 52 hours (DENV1, DENV3). Cells were fixed with 100 µL of 1:1 methanol:acetone for 30 minutes. 100 µL of permeabilization buffer was added for 10 minutes followed by 100 µL of blocking buffer (3% normal goat plasma in permeabilization buffer) and left overnight at 4°C. 50 μL of 4G2 at 12.5 ng/μL were added to the plates and incubated for 1 hour at 37°C. Cells were washed with a microplate washer followed by the addition of 50 µl of 1:3000 horseradish peroxidase-conjugated goat anti-mouse secondary antibody for 1 hour at 37°C. Foci were visualized with 100 µL of True Blue and counted with a user-supervised automated counting program on 2x-magnified images of micro-wells obtained on a CTL ELISPOT reader. NHS controls were included on every plate to define 100% infection. The eFRNT value is a discrete number corresponding to the dilution factor at which 50% maximum FFU are observed or the average of the two dilution factors between which 50% FFU is crossed.

### Real-time Polymerase Chain Reaction

Whole blood was drawn into serum serparator tubes and centrifuged. Viral RNA was extracted from 140 µL of serum using the QIAamp extraction kit (QIAGEN Hilden, Germany) and eluted in 60 µL. Real time PCR was performed on the Rotor-Gene Q (QIAGEN Hilden, Germany) platform using the RealStar ® kits for Dengue, V2.0, Chikungunya, V2.0 and Zika V.1.0 (Altona Diganostics, Hamburg, Germany) in accordance with manufacturer’s instructions.

### Statistical analysis

Data was collected in a REDCap database and analyzed with Stata 15.1 (College Station, TX) [24]. We summarized participant characteristics and compared them between those with positive and negative mRDT resuts using Student’s t-test (normally distributed data) or Wilcoxon rank-sum test (nonparametric data) for continuous variables and Pearson χ^2^ test for categorical variables. In a post-hoc analysis, we performed multivariable regression analyses to explore the demographic, geographic, and clinical parameters associated with the primary outcome of interest: a positive dengue IgG. All variables that were significant in univariate models with a pre-specified *p*-value of < 0.25 were included in the subsequent multivariate analysis [25]. A resulting *p-*value of < 0.05 was considered statistically significant in the final models.

## Results

A total of 1,702 children met the eligibility criteria and were screened for malaria with a mRDT. Complete laboratory results were available for 1,693 (99.5%) particpants, who were subsequently included in the analysis. Characteristics of the cohort are summarized in **Table 2**. The median age was 5 years (IQR 2 – 10), although children at Bugoye were significantly older (10 years, IQR 5 – 14, p< 0.0001), likely attributable to an existing village-based, community health worker led fever management intervention targeting children less than five years of age and reducing the number of these patients that presented to our study site [26].

**Table 2:**
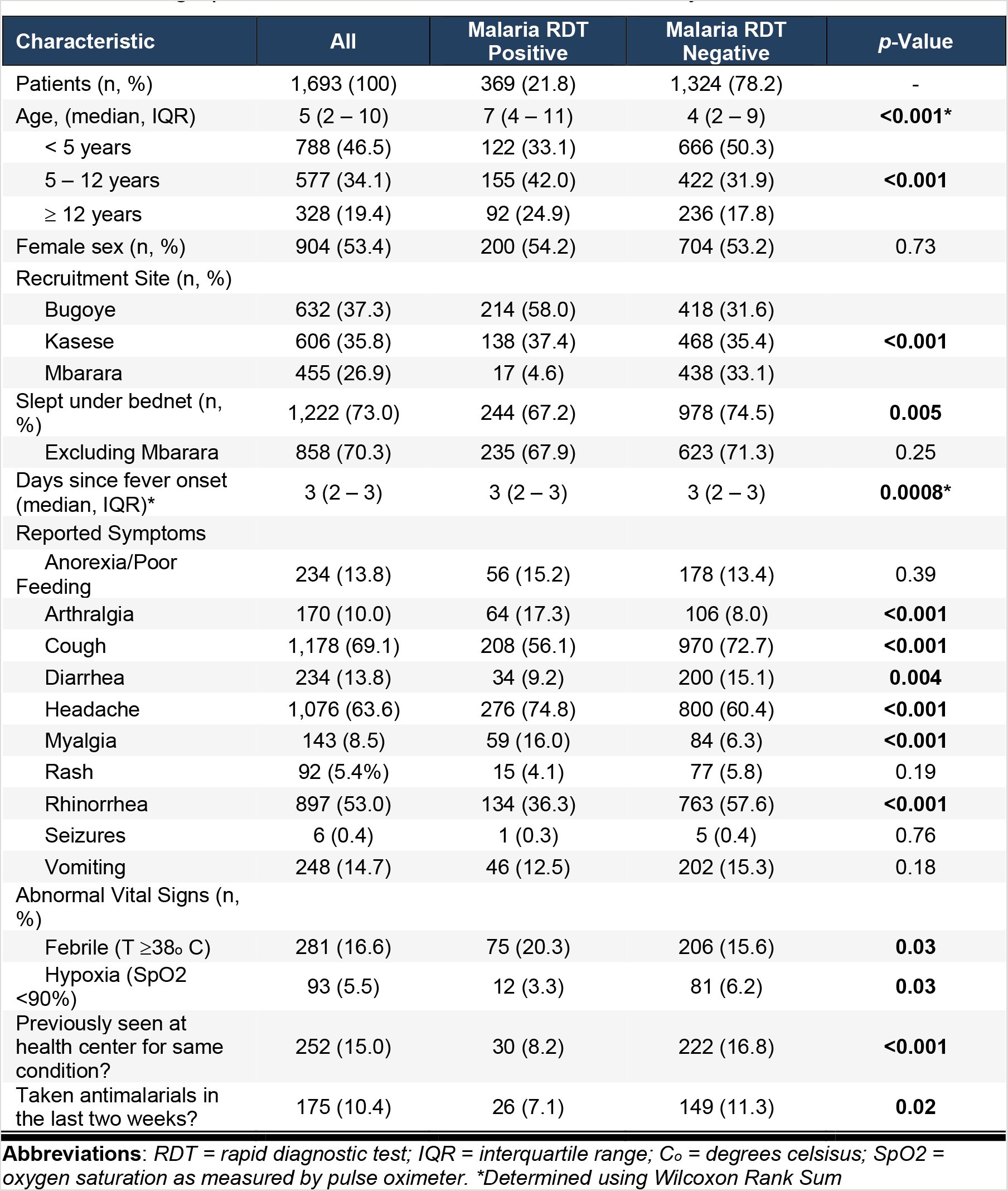
Demographic and clinical characteristics of the study cohort

Overall, 369 children (21.8%) had a positive mRDT result. The lowest rate was observed at MRRH (17 of 455, 3.7%, 95% CI 2.3 – 5.9), whereas much higher rates were seen in Kasese (138 of 606, 22.8%, 95% CI 19.6 – 26.3) and Bugoye (214 of 632, 33.9%, 95% CI 30.3 – 37.7). Self-reported ITN use was associated with a protective effect against malaria (IRR 0.70, 95% CI 0.54 – 0.90, p = 0.006), but when excluding the low-transmission site, the effect was no longer significant (IRR 0.85, 95% CI 0.65 – 1.11, p = 0.25).

There was overlap in self-reported symptoms between children with positive and negative mRDT results (**Table 2**). Among all participants, cough (69.1%), headache (63.6%), and rhinorrhea (53.0%) were the most frequently reported symptoms. A total of 431 (25.4%) participants reported all three of these symptoms, 364 (84.5%) of whom had a negative mRDT result. Participants with a negative mRDT were also more likely to have sought care and received treatment for malaria within the past 14 days. In contrast, arthralgia and myalgia were more frequently reported in the mRDT positive group.

Of the 1,324 participants with a negative mRDT test result, 1,285 (97.1%) had an evaluable dRDT result, of which 5 (0.39%) had a positive result, defined as a visible NS1, IgM, or IgG band. Individual tests results include one child with positive IgM bands, three with positive IgG bands, and one with positive IgM and IgG bands. There were three positive dRDT results in Bugoye, two in Mbarara, and and one in Kasese (**Figure 2**) All samples from MRRH tested by RT-PCR were negative for DENV, CHIKV, and ZIKV.

**Figure 2:**
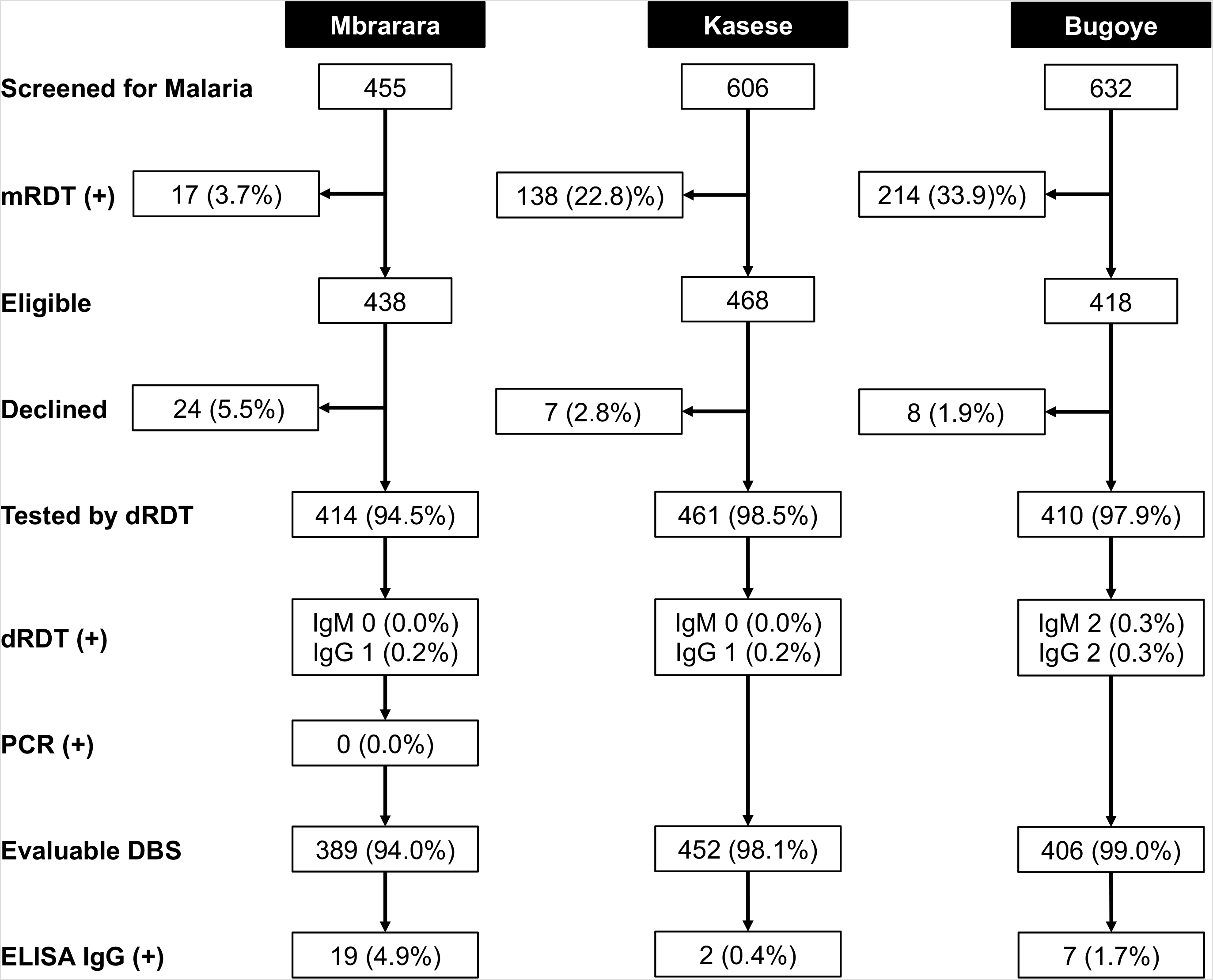
Summary of study participantion and test results stratified by study site.

Serological testing of 1,247 DBS samples from participants with a negative mRDT test result identified reactive IgG antibodies in 28 (2.3%) participants. There was a significant difference in seroprevalence between Mbarara (19 of 392, 4.9%), which is a more urban center, and the rural sites (Kasese 2 of 453, 0.4%; Bugoye 7 of 414, 1.7%; p< 0.001). The Mbarara study site was the only significant predictor of a dengue infection in the multivariable analysis when using Kasese as the reference, although there was a statistical trend towards males being at higher risk of infection (p = 0.10) (**Table 3**). Samples that were positive by RDT or IgG ELISA were also tested by eFRNT and DENV IgM-ELISA, but none were positive for IgM antibodies.

**Table 3:** Regression modeling of variables associated with presence of dengue IgG

| Variable | Unadjusted |  |  | Adjusted* |  |  |
| --- | --- | --- | --- | --- | --- | --- |
|  | OR | 95% CI | p-value | aOR | 95% CI | p-value |
| Male sex | 2.08 | 0.95 - 4.54 | 0.07 | 1.93 | 0.88 – 4.25 | 0.10 |
| Age category |  |  |  |  |  |  |
| < 5 years |  | REF |  |  |  |  |
| 5 – 12 years | 1.42 | 0.62 – 3.25 | 0.41 |  |  |  |
| ≥ 12 years | 1.10 | 0.39 – 3.16 | 0.86 |  |  |  |
| Study site |  |  |  |  |  |  |
| Kasese |  | REF |  |  | REF |  |
| Bugoye | 3.88 | 0.80 – 18.8 | 0.09 | 4.08 | 0.84 – 19.8 | 0.08 |
| Mbarara | 11.5 | 2.66 – 49.6 | 0.001 | 11.2 | 2.59 – 48.5 | **0.001** |
| Bednet use | 1.61 | 0.61 – 4.27 | 0.34 |  |  |  |

\* Variables that were significant in univariate models with a pre-specified P-value of <0.25 were included in the subsequent multivariate analysis

Of the 28 participants with a reactive IgG antibodies by ELISA, only 1 (3.6%) had a positive IgG band on the dRDT. This occurred in the one patient with both IgM and IgG bands positive on the dRDT. Overall, the correlation in IgG results between the dRDT and ELISA was poor (κ = 0.06, 95% CI 0 – 0.40).

Among those with a negative mRDT result, 1,297 of 1,324 (98.0%) received treatment with an antibiotic, including five who also received treatment with artemether/lumefantrine (AL). An additional 17 (1.3%) received treatment with AL (and no antibiotic) despite negative mRDT test results. The vast majority of children (1,279 of 1,316, 97.2%) were discharged from the outpatient clinics, while 35 (2.7%) were admitted. There were no significant differences in admission rates by sex, age category, or clinical site.

## Discussion

Despite favorable climate and ecology, our findings suggest that dengue is an uncommon cause of non-malarial, pediatric febrile illness in western Uganda. Our results are consistent with those obtained from a laboratory-based surveillance system of inpatient pediatric admissions across six sentinel sites in Uganda, which reported recent arboviral infection in only 18 of 622 (2.9%) samples tested [27], as well as multi-site seroprevalence study of healthy adult blood donors, which identified dengue antibodies in 72 of 1,744 (4.1%) of donors [28]. However, these data exclude neither the possibility of sporadic dengue outbreaks in the past or future nor the possibility that dengue or other *Aedes*-borne viruses could establish endemicity in this region given the favorable ecological conditions. Our time-limited and geographically-restricted study design, however, does provide a robust snapshot of current arbovirus transmission. While seroprevalence studies that include older subjects may increase the sensivtivity for detecting historic dengue transmission, our findings do provide strong evidence for the presence of low-level transmission, particularly at the more urban site in Mbarara. In areas of higher transmission, dengue seroprevalence is typically 20% or greater in young children [29–31]. The few cases detected here, along with the large susceptible population make dengue introduction a major concern, and surveillance together with further investigation of the demographic, spatial, and entomologic factors that could support epidemic and endemic dengue infection in urban East Africa should be a high priority.

Given the low number of positive cases, a rigorous assessment of the performance of the dengue rapid diagnostic tests was not possible. The relatively poor correlation in IgG results between the dRDT and the ELISA is consistent with a study of febrile outpatients conducted in the Democratic Republic of Congo, which showed 2.5% IgG positivity by dRDT and 34.0% positivity by ELISA with an estimated sensitivity of 7.6% [32]. It is crucial to note that dRDT are designed for use in the setting of acute infection and not for seroprevalence studies. The main advantage of including IgG testing in this context is in secondary infection, when IgG may rise quickly and dramatically, suppressing the IgM response and limiting unbound dengue antigen present in serum [33]. In primary infection, patients would typically present before an IgG response has developed. A recent systematic review evaluated the potential of dRDT to function as a measure of seroprevalence but found performance to be suboptimal, with sensitivity ranging from 30–60% [34].

Our results do highlight the urgent need for better diagnostics and management algorithms for children with acute febrile illness. Malaria was identified as the cause of fever in only 22% of participants, and among participants with a negative malaria test, nearly 80% reported cough or rhinorrhea with more than half (51.1%) reporting both symptoms. While not confirmed by microbiologic or molecular methods, many of these children likely had viral infections that would be self-limited and not require antibiotic treatment [35]. Yet more than 97% of children with a negative malaria test result across all three sites received a course of antibiotic treatment. Given the growing global threat of antimicrobial resistance, driven in part by antimicrobial overuse, this patient population represents an important target for antibiotic stewardship interventions.

To our knowledge, our study is one of the first to examine the burden of dengue among children presenting to outpatient clinics in western Uganda with undifferentiated febrile illness. Strengths of our approach include the diverse geography of the clinical sites, large sample size, and different diagnostic testing strategies (i.e. dRDT, ELISA, PCR). The study also has a number of limitations. First, we excluded children with a positive m RDT from further consideration. While dengue-malaria coninfections have been reported in other settings [6], this was expected to be a relatively infrequent occurrence. To explore this possibility, however, we tested 131 mRDT positive children at the Kasese site, only one (0.77%) of who had a positive IgM band on the dRDT; a prevalence similar to children with a negative mRDT (p = 0.53). Second, our facility-based and temporally-limited enrollment strategy may have missed focal outbreaks of dengue in the community. Dengue is well-known to emerge rapidly in non-immune populations then quickly disappear. Studies from neighboring countries and other sureveillance methods do, however, support he validity of our findings. Third, IgG ELISA may overestimate dengue prevalence as this assay may also detect cross-reactive antibodies elicited by infection by related flaviviruses, though neutralization testing also supported prior infection by dengue for some subjects in this population [36, 37]. Lastly, we did not have the means to screen all specimens for anti-dengue IgM, which precluded a direct comparison of serologic IgM testing to the dRDT result.

## Conclusions

Dengue does not account for a large proportion of non-malarial, acute febrile illness presentations in western Uganda. However, the presence of the vector and the pathogen, along with a large susceptible population make sporadic outbreaks a major concern. Further investigation into the eocological factors that sustain low-level transmission in urban centers such as our Mbarara site, are urgently needed.

## Data Availability

Deidentified individual data that supports the results will be shared beginning 9 to 36 months following publication provided the investigator who proposes to use the data has approval from an Institutional Review Board (IRB), Independent Ethics Committee (IEC), or Research Ethics Board (REB), as applicable, and executes a data use/sharing agreement with UNC.

## Abbreviations

ACT: Artemisinin combination therapies
AL: Artemether/lumefantrine
C°: Degress celsius
CHIKV: Cikunguunya Virus
DBS: Dried Blood Spot
DENV: Dengue Virus
dRDT: Dengue rapid diagnostic test
ELISA: Enzyme Linked immunosorbent Assay
ITN: Insecticide-treated nets
mRDT: Malaria rapid diagnostic test
MRRH: Mbarara Regional Referral Hospital
NS1: Non-structural protein 1
OD: Optical Density
RNA: Ribonucleic acid
RT-PCR: Reverse-Transcriptase Polymerase Chain Reaction
ZIKV: Zika Virus

## Declarations

## Ethics Approval and Consent to Participate

Ethical approval of the study was provided by the institutional review boards of the University of North Carolina at Chapel Hill, the Mbarara University of Science and Technology, and the Uganda National Council for Science and Technology. Written informed consent for participation in the study was obtained from all adult participants or from their parent or guardian when participants were children (under 18 years old).

## Consent for Publication

Not applicable.

## Author Contributions

Study conception and design: RMB, MHC, EM. Funding: RMB. Study implementation: RMB, RM, RK, EC MM, MN, DN, EM. Laboratory testing: RK, DN, SG, DE, YZ, MHC. First draft of manuscript: RMB, SG, EC, MHC, DN. Revisions: All.

## Acknowledgements

We wish to thank the clinical staff and patients of the Bugoye Health Centre, Kasese Health Centre, and Mbarara Regional Referral Hospital Outpatient Department for their participation and support. We also thank Corinna Keeler for the cartography support.

